# U-shaped Association of Non-HDL-C/HDL-C Ratio with Atrial Fibrillation Recurrence Post-Catheter Ablation: A Cohort Study

**DOI:** 10.1101/2025.05.30.25328684

**Authors:** Mingjie Lin, Hao Wang, Nan Yang, Tongshuai Chen, Bing Rong, Wenqiang Han, Jingquan Zhong, Kai Zhang

## Abstract

**Background:** The prognostic value of lipid metrics in atrial fibrillation (AF) recurrence following catheter ablation (CA) remains controversial. This study investigated the effectiveness of novel lipid parameter non-high-density lipoprotein-cholesterol (HDL-C)/HDL-C ratio (NHHR) in predicting post-ablation AF recurrence.

**Methods:** In this cohort study, 1,728 patients with AF undergoing first-time CA at our center (2017– 2020) were stratified by NHHR quartiles (Q1–Q4). The primary outcome was AF recurrence following CA. Logistic regression analysis was used to detect the association between NHHR and major adverse cardio- and cerebrovascular events (MACCEs).

**Results:** A U-shaped relationship was observed between the NHHR and AF recurrence (P_nonlinear_=0.021), with an inflection point at NHHR=2.61. Compared to the optimal range (2.62-3.26), patients with Q1 of NHHR had a 63.6% higher recurrence risk (hazard ratio [HR]=1.636, 95% confidence interval [CI]: 1.235–2.166). In cases with the NHHR < 2.61, the AF recurrence risk decreased with an adjusted HR of 0.715 (95% CI, 0.524–0.977, P = 0.035) for every one unit increment in the NHHR. Subgroup and sensitivity analyses confirmed consistency. NHHR demonstrated no association with MACCEs, underscoring its AF-specific prognostic value.

**Conclusions:** This study demonstrated a novel U-shaped association between NHHR and AF recurrence following CA. NHHR serves as a robust biomarker for AF recurrence risk stratification post-ablation, reflecting bidirectional lipid pathophysiology beyond atherosclerotic pathways. These findings advocate for NHHR integration into clinical monitoring protocols while cautioning against therapeutic NHHR manipulation without mechanistic validation.

**Trial registration:** ChiCTR-OCH-14004674

## BACKGROUND

Atrial fibrillation (AF), which is the most common type of arrhythmia in clinical practice, affects nearly 52.6 million people worldwide [1, 2]. AF causes various complications, including ischemic stroke, heart failure, myocardial infarction, and even cardiovascular mortality; moreover, AF associated socioeconomic and healthcare costs are expected to increase in the foreseeable future [3]. Catheter ablation (CA) has emerged as a cornerstone intervention for symptomatic AF refractory to pharmacotherapy; CA is endorsed by current ESC guidelines as a Class I recommendation for paroxysmal AF (PAF) and certain persistent AF cases [1, 4, 5]. However, the overall success rate of CA for AF is not ideal, and the relatively high risk for tachycardia recurrence is concerning. Thus, the reduction of the AF recurrence rate following CA is an important clinical issue. Hence, identifying the particular characteristics of patients with recurrence following CA is crucial.

The complex interplay between dyslipidemia and AF pathogenesis has garnered increasing attention, particularly regarding the “cholesterol paradox” phenomenon stating that high total cholesterol (TC) or low-density lipoprotein (LDL) was associated with a lower risk of AF prevalence [6, 7]. A meta-analysis, however, revealed that higher TC levels were independently attributed to an increased risk of AF in individuals without cardiovascular disease (CVD) [8]. Although few studies have evaluated on the relationship between blood lipids and AF recurrence following CA, the results were not entirely consistent. A previous study demonstrated the LDL-C levels to be negatively associated with the AF recurrence risk following CA [9], while another study demonstrated elevated triglycerides (TG) levels measured before CA to be associated with an increased AF recurrence risk [10]. These discrepancies underscore the limitations of isolated lipid fraction analysis and highlight the need for composite indices integrating atherogenic and protective lipid components.

The non-high-density lipoprotein-cholesterol (HDL-C)-to-HDL-C ratio (NHHR), a novel compound lipid parameter, synergistically encapsulates atherogenic lipid burden (non-HDL-C: LDL-C + remnant cholesterol) and reverse cholesterol transport capacity (HDL-C) [11]. Recent studies have validated a U-shaped association between NHHR and major cardiovascular events [12, 13]. Nevertheless, the prognostic value of NHHR in post-ablation AF recurrence remains unexplored, representing a critical knowledge gap in electrophysiological risk stratification. To address this gap, we designed this study utilizing data gathered at the Qilu Hospital of Shandong University from 2017 to 2020 to investigate the association between NHHR and AF recurrence in patients undergoing CA.

## METHODS

### Study participants and ethics approval

This retrospective cohort study consecutively enrolled patients with AF aged 18–80 years undergoing first-time CA at the Qilu Hospital of Shandong University between January 2017 and December 2020. Exclusion criteria included valvular heart disease[14], follow-up duration <2 years, or incomplete lipid profiles. Ethical approval was obtained from the Institutional Review Board of the Qilu Hospital (Approval No. [2014]008), adhering to the Declaration of Helsinki principles. Written informed consent was obtained from all participants, with data sourced from the registered prospective study (ChiCTR-OCH-14004674) and managed in compliance with anonymization protocols.

### Baseline data collection and definitions

Baseline parameters were systematically extracted from electronic medical records using a standardized case report form. Variables included demographics (sex, age, body mass index [BMI[), AF subtype classification (paroxysmal vs. persistent per 2020 ESC guidelines [15]), comorbidities (congestive heart failure, hypertension, diabetes, and prior stroke/transient ischemic attack/systemic embolism, vascular disease), CHA_2_DS_2_-VASc and HAS-BLED scores [11], and lipid profiles (TC, TG, LDL-C, HDL-C). Non-HDL-C was calculated as TC−HDL-C, with NHHR defined as HDL-C/Non-HDL-C [11]. Other data included left atrial diameter (LAD), agents (anti-arrythmic drugs, anticoagulants, antiplatelet drugs, angiotensin-converting enzyme inhibitor/ angiotensin receptor blockers, and statins).

### CA procedure

Before the ablation procedure, transesophageal echocardiography and/or contrast-enhanced computed tomography were performed to exclude the left atrial (LA) thrombus and clarify the anatomy of the LA and pulmonary veins (PV). Oral anticoagulant drugs were administered before admission or immediately after admission, and were used continuously during the perioperative period. Antiarrhythmic drugs other than amiodarone, were discontinued use for five half-lives.

The method of CA, i.e., radiofrequency ablation (RFA) or cryoablation, was selected based on the discussion between the patient and physician.

RFA was performed using a three-dimensional mapping system (CARTO 3, Johnson & Johnson Inc., USA). During the surgical process, Lasso or PentaRay mapping catheter was used to reconstruct the three-dimensional model of the LA, and then RFA was performed by an open-irrigated tip catheter with contact force sensing (typically set to upper-temperature limit of 43 °C, power of 35–45 W, and an infusion rate of 20–25 ml/min). PV isolation (PVI) was performed by experienced operators, and additional CA procedures, including LA roof line, LA posterior wall line, mitral valve isthmus, tricuspid valve isthmus, and superior vena cava or substrate modification under sinus rhythm, were decided by the operators. If AF persisted after CA completion, electrical conversion was performed to restore the sinus rhythm.

Cryoablations were performed to achieve PVI in selected patients with PAF by using a 28-mm second-generation cryoballoon catheter (Arctic Front Advance, Medtronic Inc., USA). Cryoablation entailed applications lasting 120–240 s in each PV was repeatedly applied while monitoring PV potentials with the circular mapping catheter (Achieve, Medtronic Inc., USA). If PVI was not achieved after repetitive cryoablations, a supplementary ablation was added using a radiofrequency catheter.

### Postoperative management and follow-up

After CA, all patients took anticoagulants and proton pump inhibitors for 3 months. The use of antiarrhythmic drugs was determined by clinical doctors and continued for up to 3 months following CA. After 3 months, anticoagulant treatment continuation was determined based on the CHA_2_DS_2_-VA score. Electrocardiogram (ECG) and 24-h Holter monitoring were conducted at 3, 6, and 12 months of the first year after intervention. Additionally, after 1 year of ablation, telephonic follow-up was performed every 6 months to report any discomfort; patients were recommended to undergo an ECG and contact our study team in case of the development of cardiac symptoms. The primary endpoint was AF recurrence, defined as any atrial tachyarrhythmia lasting >30 s after the blanking period. Major Adverse Cardiac and Cerebrovascular Events (MACCE) are defined as cardiovascular death (e.g., fatal myocardial infarction, sudden cardiac death), myocardial infarction (both non-fatal and fatal events), stroke (ischemic or hemorrhagic); revascularization (repeat percutaneous coronary intervention or coronary artery bypass grafting), and hospitalization for unstable angina or heart failure.

### Statistical analysis

Continuous variables are expressed as mean ± standard deviation (SD) or median (interquartile range) based on normality assessed by the Shapiro–Wilk test. Categorical variables are reported as frequencies (percentages). Between-group differences across NHHR quartiles were analyzed using one-way analysis of variance (ANOVA) (normally distributed data), Kruskal–Wallis test (non-normal data), or chi-square test (categorical variables). Cox regression models were used to investigate whether there was a trend between ordered NHHR quartiles and AF recurrence with NNHR quartile ranks used as continuous variable. AF-free survival was evaluated by Kaplan–Meier curves with log-rank tests stratified by NHHR quartiles. Cox proportional hazards models were used to estimate hazard ratios (HRs) and 95% confidence intervals (CIs) for AF recurrence. All baseline variables (demographics, comorbidities, laboratory parameters, and medications) were initially screened in the univariable analysis. Variables with P < 0.05 in the univariable analysis or clinical relevance (e.g., age, sex, left atrial diameter [LAD], diabetes mellitus) were included in a forward stepwise Cox model.

Restricted Cubic Spline (RCS) Analysis: Nonlinear relationships between NHHR and AF recurrence were explored using RCS with five knots. The likelihood ratio test compared the linear and nonlinear models. A U-shaped association was confirmed if the nonlinear term achieved P < 0.05. The inflection point was identified via recursive algorithms. Prespecified subgroups (age, sex, AF type, CHA_2_DS_2_-VA score, LAD, non-PVI lesions, and diabetes) were analyzed using stratified Cox models. Interaction terms (NHHR quartile × subgroup variable) were tested via Wald tests with Bonferroni correction for multiplicity (P < 0.05/7). MACCEs were analyzed via logistic regression due to low event rates. Variables with P < 0.05 in the univariable analysis were included in the multivariable models. Analyses were performed using R v4.2.3 (R Foundation) and SPSS v26.0 (IBM). Two-sided *P* < 0.05 indicated statistical significance.

Sensitivity Analyses: Temporal Robustness: Primary models were reanalyzed using the 24-month follow-up data. Outlier Exclusion: NHHR values beyond ±2 SD were excluded to mitigate extreme-value effects.

## RESULTS

### Baseline characteristics

A total of 1914 patients with AF who underwent CA were screened for eligibility and ultimately 1728 patients were included in the final analysis (Fig. 1). The cohort exhibited a mean age of 60.22 ± 10.55 years (62.3% male), with an average follow-up duration of 42.99 ± 13.24 months (Table 1). Notably, no interquartile disparity in the follow-up duration was observed among the NHHR quartiles (*P*=0.719). A clear inverse relationship was observed between the NHHR quartiles and age (*P* < 0.001). Male prevalence increased progressively across the quartiles (*P* < 0.001). Hypertension prevalence declined from Q1 (54.7%) to Q3 (44.4%) but rebounded in Q4 (49.0%, *P* = 0.023), while vascular disease revealed a stepwise reduction (Q1: 28.7% vs. Q4: 13.6%, *P* < 0.001). BMI and lipid profiles demonstrated strong positive associations with NHHR quartiles. BMI escalated from 25.27 ± 3.35 kg/m² (Q1) to 27.27 ± 3.36 kg/m² (Q4, *P* < 0.001). Similarly, TC, TG, and LDL-C exhibited significant upward trends from Q1 to Q4 (all *P* < 0.001). Conversely, the HDL-C levels declined progressively (*P* < 0.001). The CHA_2_DS_2_-VA and HAS-BLED scores decreased with higher NHHR quartiles (both *P*< 0.001). Antiarrhythmic drug usage declined in Q4 (6.2% vs. Q1: 11.0%, *P* = 0.007), whereas statins utilization decreased from 51.9% (Q1) to 38.1% (Q4, *P* < 0.001).

**Fig. 1.**
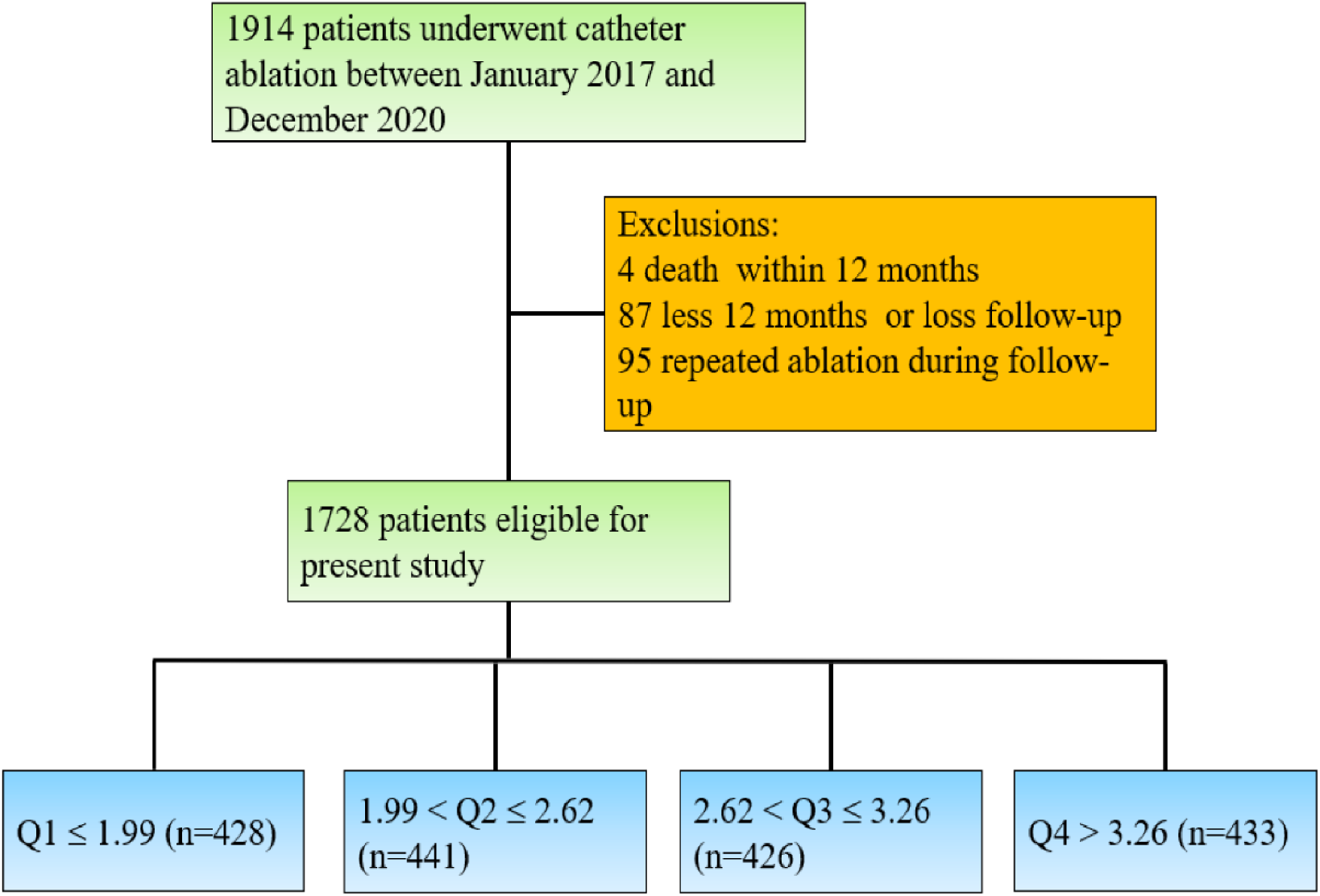
Study Design and Analytical Workflow. Schematic overview of the research methodology for investigating the association between NHHR and AF recurrence post-catheter ablation.

**Table 1.**
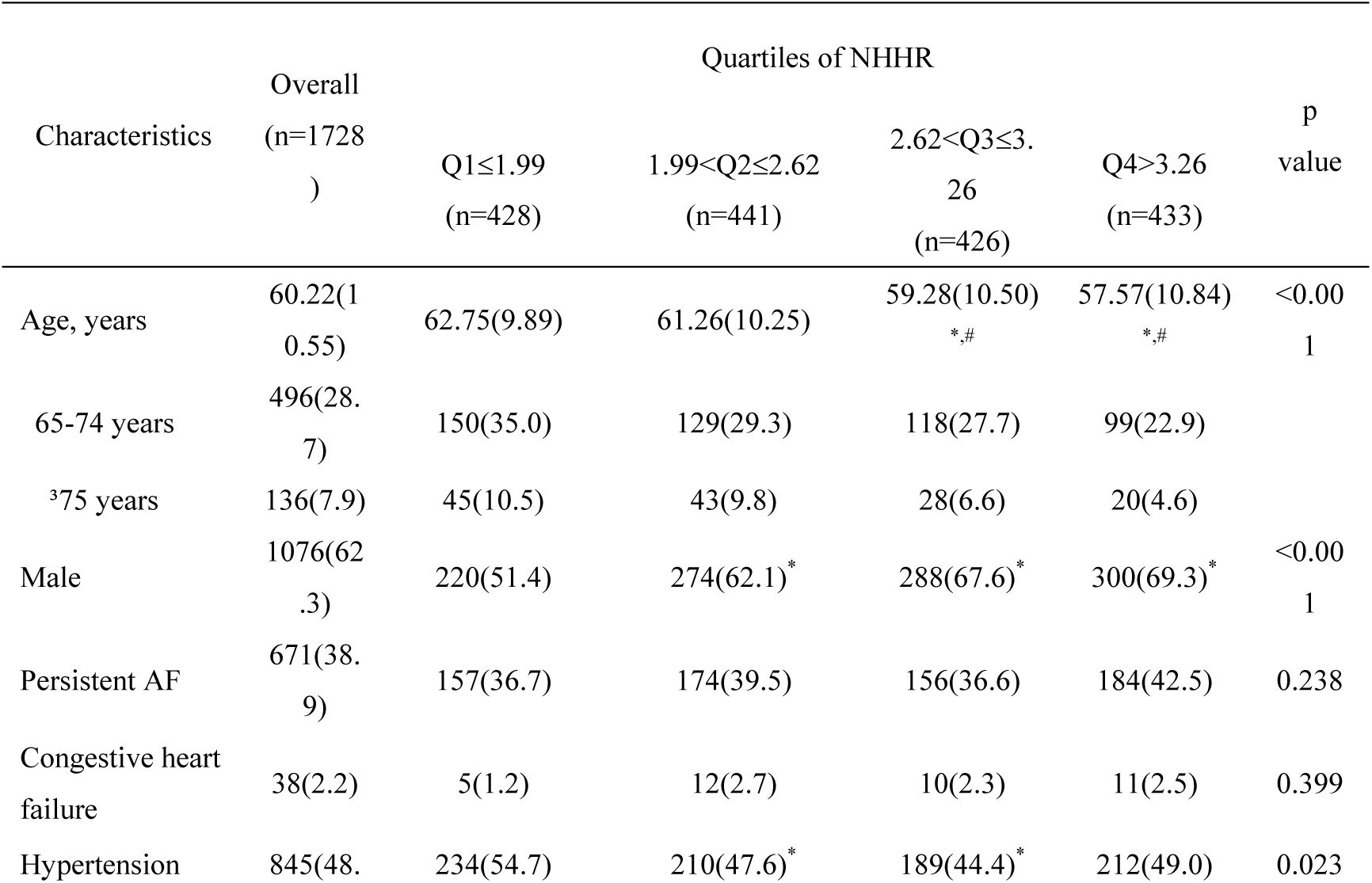
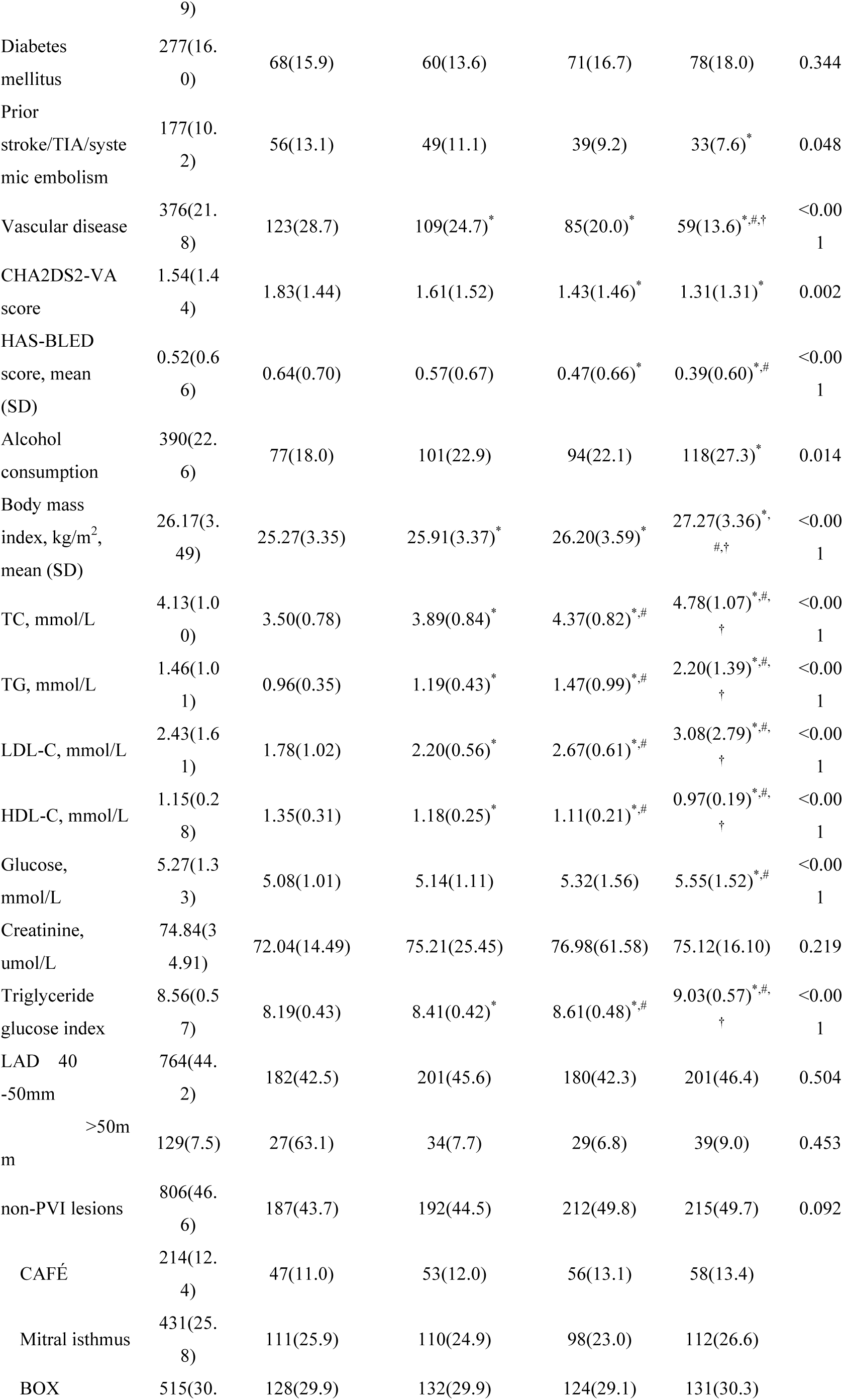
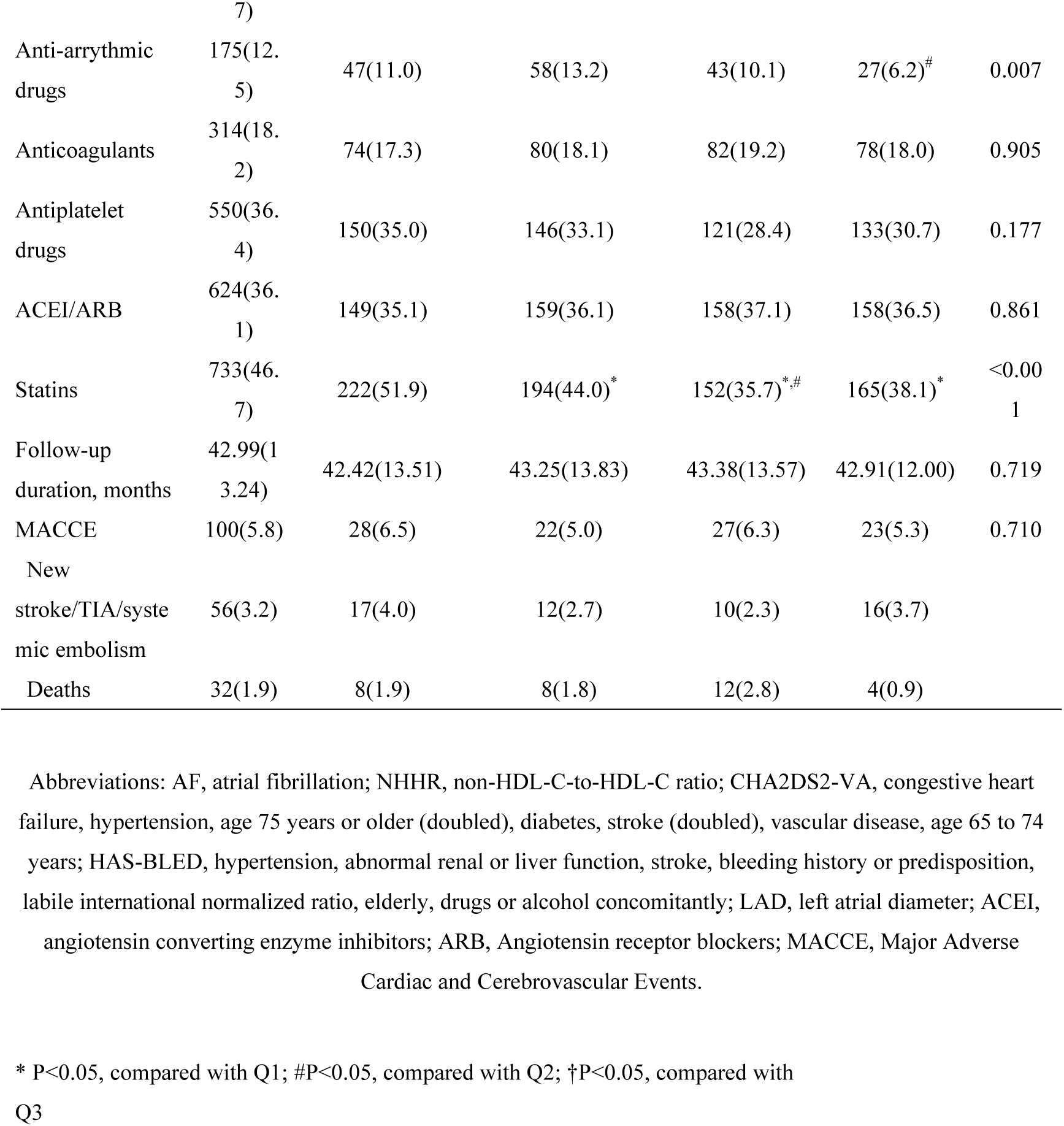
Baseline and follow-up information.

### Kaplan–Meier analysis of the AF-free survival across the NHHR groups

A total of 402 participants were recorded with AF recurrence; the AF recurrence rate was 23.3%. As shown in Fig. 2, the AF-free survival rates exhibited significant differences among the NHHR quartiles (*P* for trend = 0.004). NHHR quartile Q1 (≤1.99) demonstrated a 63.6% higher risk of AF recurrence compared to those in the third quartile (2.62–3.26) (HR=1.636, 95% CI: 1.235–2.166; P < 0.001). Stratified analyses were performed to account for potential confounding factors, such as age and sex disparities across groups. Notably, the trend in AF-free survival among the NHHR quartiles persisted for younger patients (<65 years) and male participants (*P* for trend=0.002 and 0.010, respectively; Additional files 1 and 2). Specifically, the HR of Q1/Q3 for AF recurrence in these subgroups were 2.063 (95% CI: 1.403–3.034; *P* < 0.001) for younger patients and 1.946 (95% CI: 1.308–2.896; *P* = 0.001) for males.

**Fig. 2.**
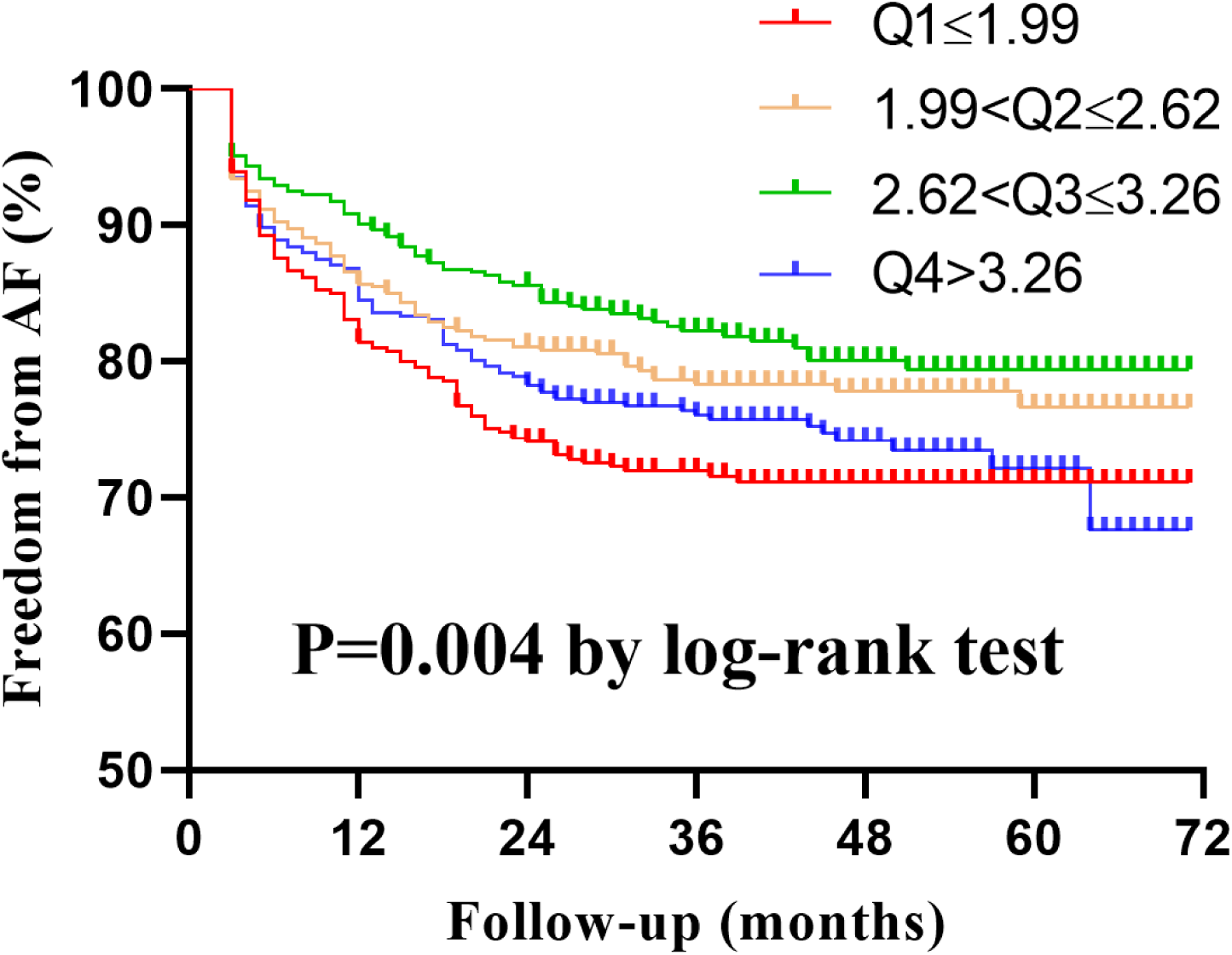
Kaplan-Meier Survival Analysis Stratified by NHHR Quartiles. Long-term AF-free survival probabilities in patients categorized by baseline NHHR levels.

### Risk factors of AF recurrence

The relationships between the several factors and AF recurrence are shown in Table 2 based on the univariate and multivariate COX proportional regression survival models. After multivariate adjustment, independent predictors of AF recurrence included risk factors (female sex [HR 1.285; 95% CI: 1.048–1.577], diabetes mellitus [HR 1.605; 95% CI: 1.262–2.042], and LAD >50 mm [HR 2.173; 95% CI: 1.575–2.997]) and protective factors (second [HR 0.736; 95% CI: 0.561–0.966] and third [HR 0.622; 95% CI: 0.467–0.829] NHHR quartiles), all statistically significant (P < 0.05). In the univariable cox analysis, we also compared the relationship between different blood lipid indicators (TC, TG, LDL-C, HDL-C, triglyceride-glucose index, and NHHR) and AF recurrence, respectively, and found that NHHR is the only one factor associated with AF recurrence. After adjusting for the factors, the third quartile demonstrated the lowest risk of AF recurrence with a statistically significant trend (*P* for trend=0.001).

**Table 2.**
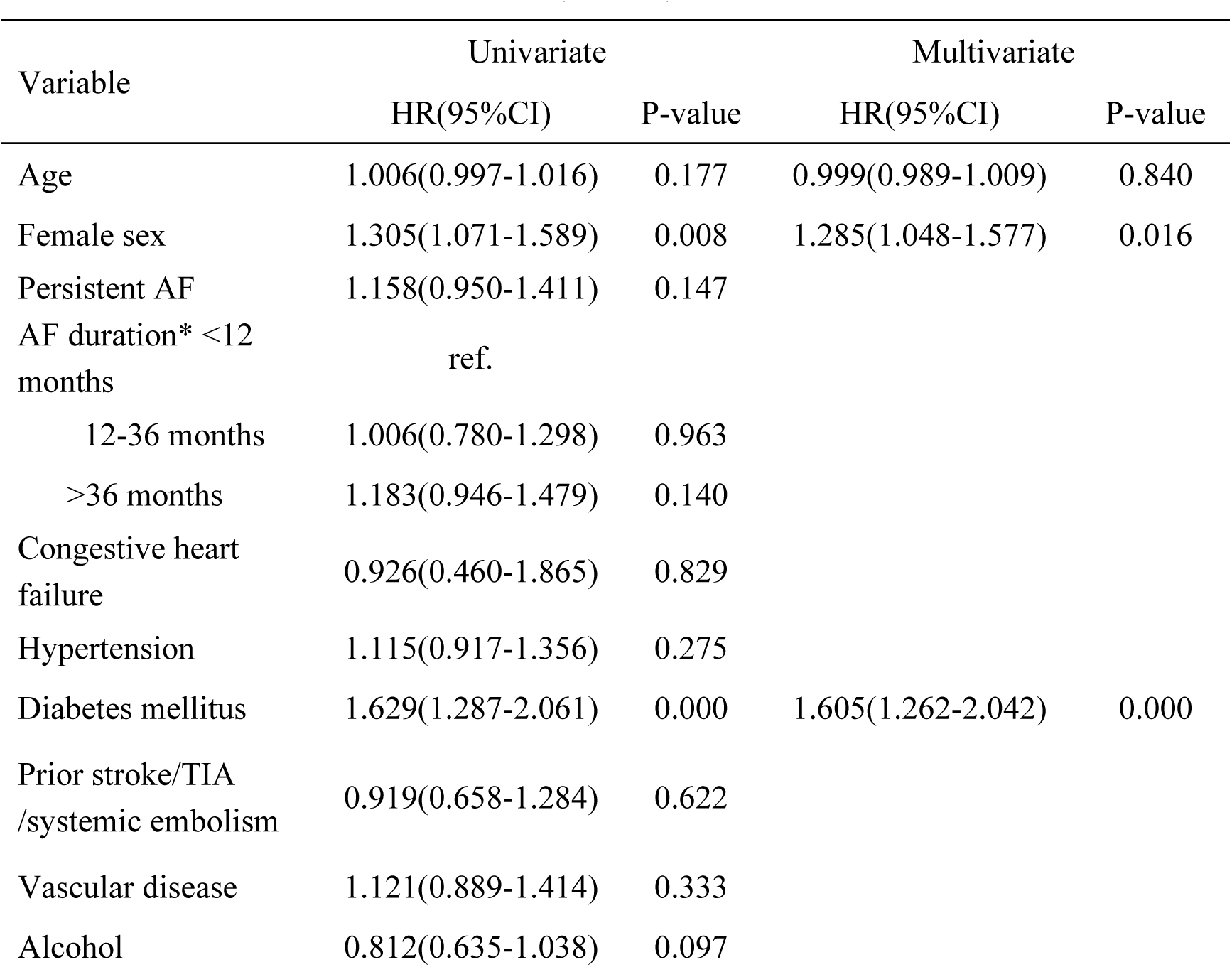
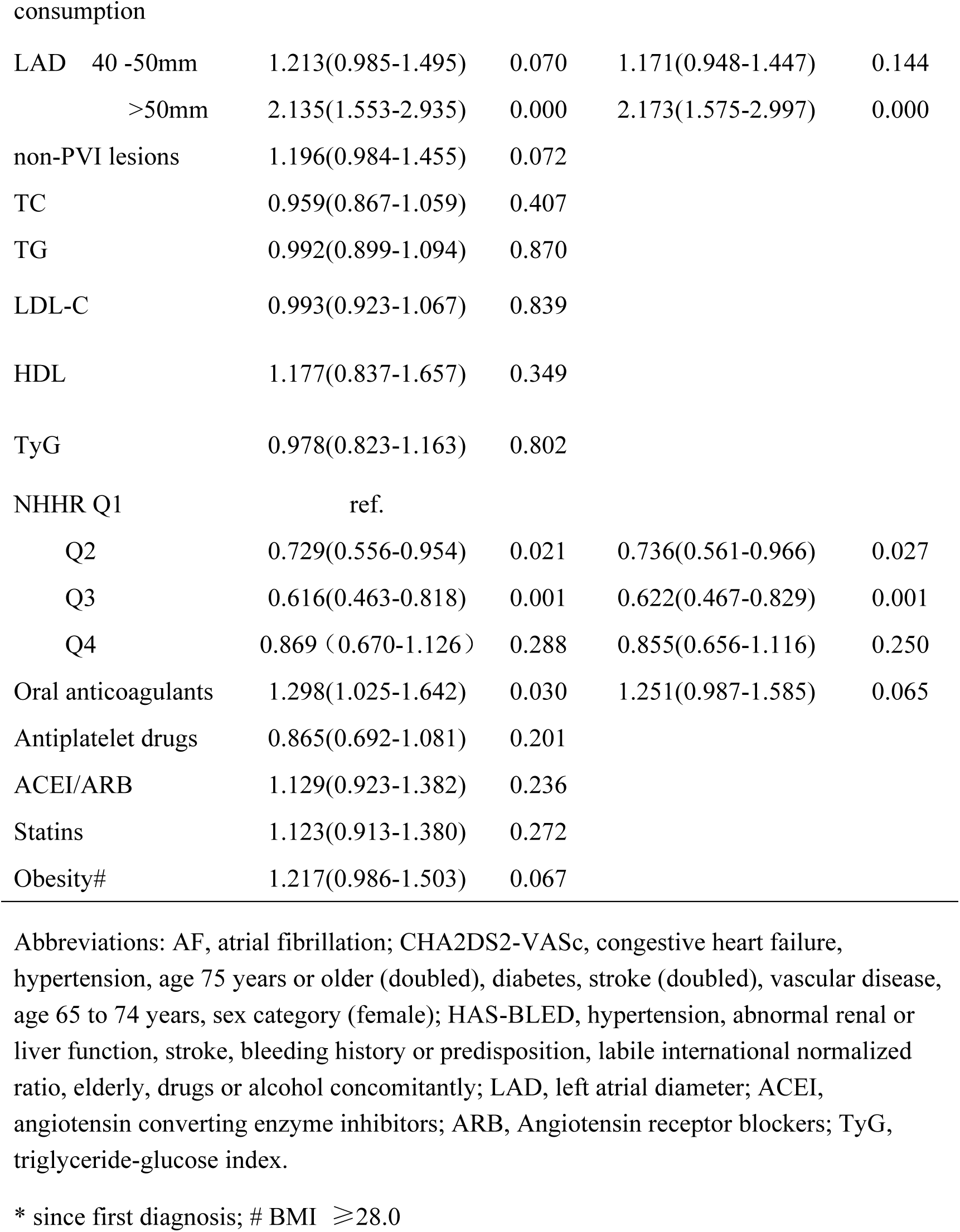
Univariable and multivariable cox regression for atrial fibrillation recurrence (n=1728)

### U-shaped relationship between NHHR and AF recurrence

In order to investigate the potential nonlinearity, we performed RCS fitted for Cox proportional hazards models to further investigate the relationship between NHHR and AF recurrence. As shown in Fig 3, a significant statistical correlation was observed between NHHR and AF recurrence (*P*=0.022) and a U-shaped association was observed between the NHHR and AF recurrence (*P* for nonlinear =0.021). After applying a recursive algorithm, the inflection point for NHHR in relation to the risk of AF recurrence was 2.61. In cases where the NHHR was <2.61, the AF recurrence risk decreased with an adjusted HR of 0.715 (95% CI: 0.524–0.977, *P* = 0.035 for every one unit increment in the NHHR. When the NHHR ≥2.61, no significant dose-effective relationship was observed (an adjusted HR of 1.102 [95% CI: 0.953–1.274]; *P* = 0.191).

**Fig. 3.**
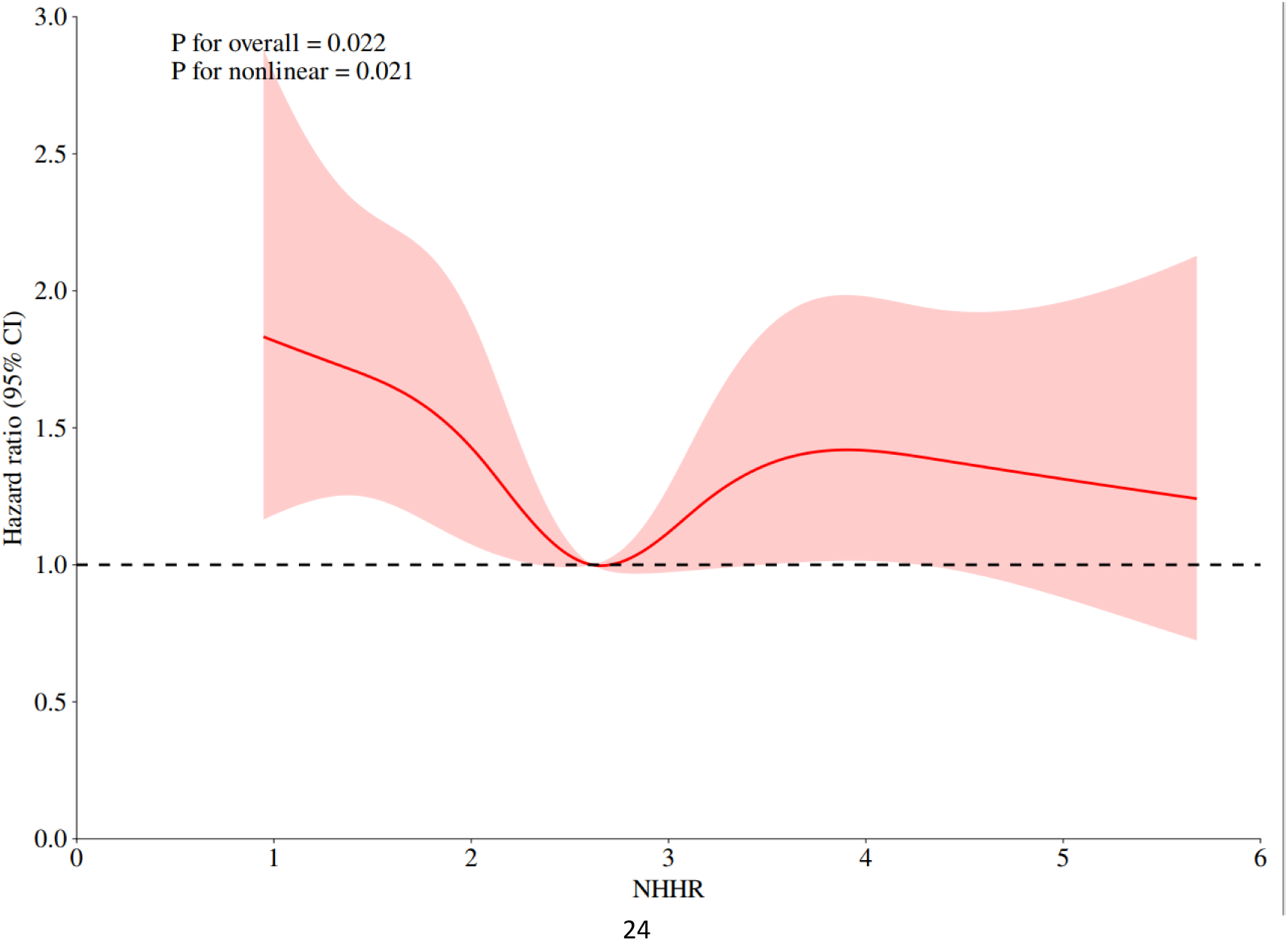
Nonlinear Association Between NHHR and AF Recurrence Risk. U-shaped association with inflection point at NHHR=2.61 with Restricted cubic spline analysis.

### Subgroup analyses

Stratified and interaction analyses were conducted to further elucidate the relationship between NHHR and AF recurrence in a series of subgroup analyses, including age (≤65 years vs. >65 years), sex, AF type (PAF vs. Persistent AF), CHA_2_DS_2_-VA score (<2 vs. ≥2), LAD (≤40 mm vs. > 40 mm), non-PVI lesions, and diabetes mellitus as detailed in Table 3. The association between NHHR and AF recurrence was consistent across all subgroups, and no significant interaction was observed between the baseline NHHR and the stratified variables (*P* for interaction > 0.05/7).

**Table 3.**
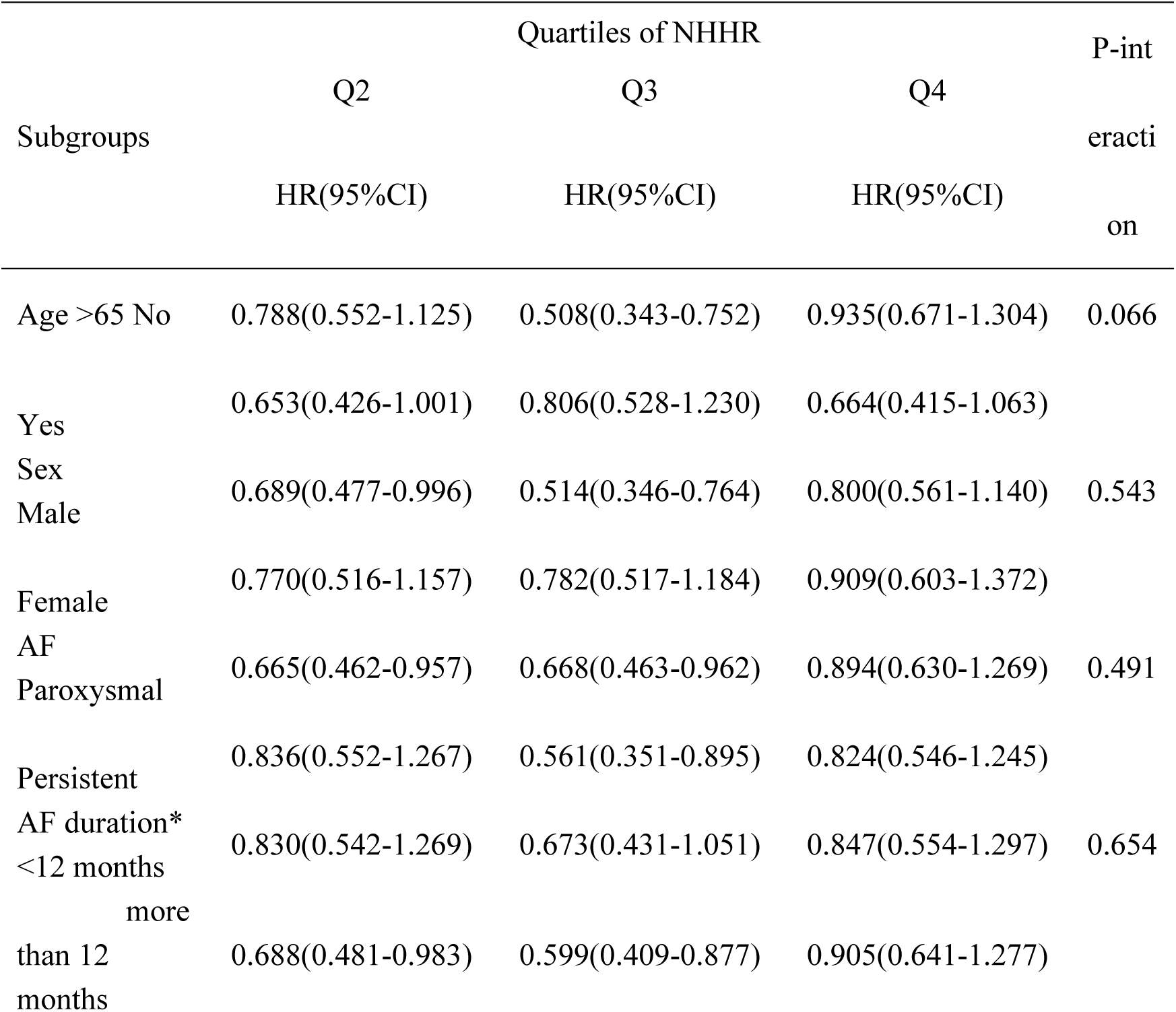
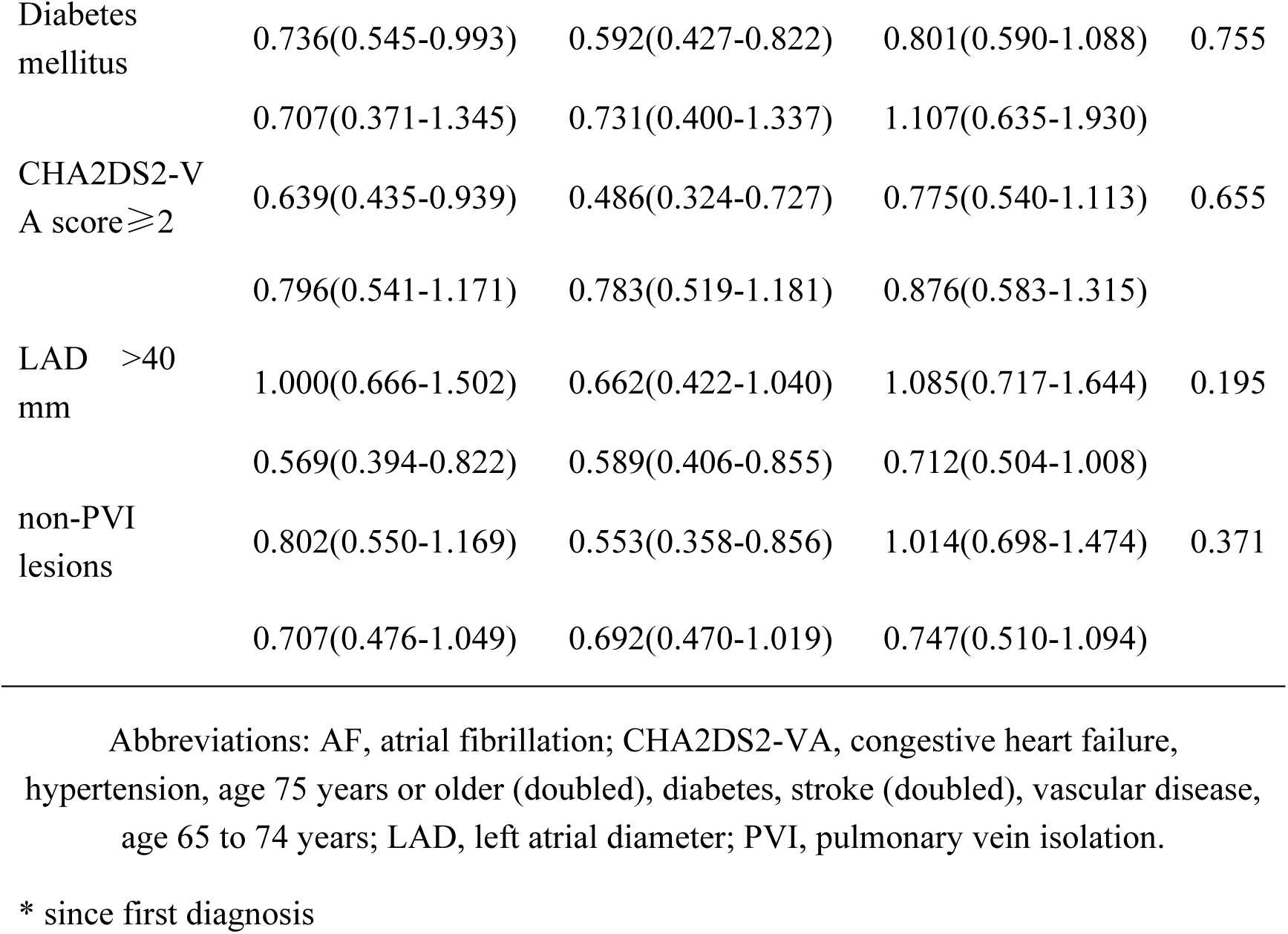
Subgroup analysis of the associations between NHHR and atrial fibrillation recurrence.

### Sensitivity analysis

To validate the temporal robustness of associations, we extended the sensitivity analyses to the 24 months follow-up duration (Additional File 3). In the unadjusted Model 1, a significant non-linear trend was observed between NHHR quartiles and risk for AF recurrence (*P* = 0.024), with the third quartile (Q3: NHHR 2.62-3.26) exhibiting the most pronounced risk reduction (HR = 0.564; 95% CI: 0.418 – 0.761). After adjusting for age, sex, diabetes mellitus, left atrial diameter, and oral anticoagulants (Model 2), the risk reduction in Q3 remained statistically significant (HR = 0.577, 95% CI: 0.426–0.781); however, the overall trend weakened (*P* = 0.050). To address potential outliers, a secondary sensitivity analysis excluded the NHHR values beyond ±2 SD (Additional File 4). The protective effect in Q3 persisted (HR = 0.614, 95% CI: 0.462–0.816), while Q4 also demonstrated attenuated risk reduction (HR = 0.879, 95% CI: 0.673–1.148) in the unadjusted model. Q3 maintained significance (HR = 0.619, 95% CI: 0.464–0.824), but Q4’s effect remained non-significant (HR = 0.872, 95% CI: 0.664–1.148) in the adjusted Model.

### Relationship between NHHR and MACCEs

During follow up, a total of 100 patients (5.8%) experienced MACCEs (Table 1 and Additional File 5). Number of MACCEs in quartiles 1 to 4 were as follows: 28 (6.5%), 22 (5.0%), 27 (6.3%), and 23 (5.3%), respectively. Univariable and multivariable logistic regression were used to evaluate the association between the several factors and MACCEs. The results demonstrated that age, diabetes mellitus, LAD (>50 mm) and oral anticoagulants were the independent risk factors for MACCEs (all *P* < 0.05). We also detected the association between lipid indicators and MACCEs. Univariable logistic regression model revealed no significant correlation between single lipid indicators (TC, TG, HDL-C and LDL-C) or comprehensive lipid index (TyG and NHHR) and MACCE events (all *P*>0.05); thus, we did not include lipid indicators in the multivariable logistic regression analysis.

## DISCUSSION

Our study firstly demonstrated a U-shaped relationship between NHHR and AF recurrence following CA, with a critical inflection point at NHHR=2.61. This extends the "cholesterol paradox" observed in AF pathogenesis to the post-ablation setting, where both extremes of lipid imbalance—excessive atherogenic burden (NHHR≥2.61) and dysfunctional reverse cholesterol transport (NHHR<2.61) predispose to arrhythmia recurrence.

The relationship between lipid profiles and atherosclerotic CVD has been extensively documented [16]. Emerging evidences have suggested that dyslipidemia may significantly influence AF risk through complex mechanisms [17, 18]. An epidemiological study demonstrated a paradoxical inverse association where the elevated TC and LDL-C levels correlated with reduced AF incidence within the initial 5-year follow-up [19]. This "cholesterol paradox" persists in post-ablation settings, as demonstrated by a Chinese cohort study which demonstrated an L-shaped relationship between preoperative LDL-C and AF recurrence, with elevated risks of AF recurrence in both lower and higher LDL-C levels compared to the 3.20 mmol/L [9]. Notably, a meta-analysis revealed no significant differences in conventional lipid markers (TC, LDL-C, HDL-C, and TG) between recurrence and non-recurrence groups following AF ablation [20], highlighting the limitations of isolated LDL-C measurements in capturing the intricate balance between atherogenic burden and protective lipid dynamics crucial for AF substrate formation. The discordant findings underscored the need to reconsider lipid metrics, as traditional parameters failed to reflect functional aspects such as reverse cholesterol transport efficiency and lipoprotein particle characteristics that may differentially impact electrical and structural remodeling in atria [21]. These observations suggested that the pathophysiological interplay between lipid metabolism and AF involves multifaceted mechanisms beyond simple cholesterol deposition, potentially including membrane cholesterol stabilization effects on ion channels, inflammatory modulation through oxidized LDL particles, and mitochondrial dysfunction induced by lipotoxicity. The clinical implications warrant investigation into composite biomarkers, such as NHHR, which incorporates both atherogenic and anti-atherogenic lipoproteins, demonstrates superior predictive capability compared to either fraction alone concerning cardiovascular-related events [22, 23]. However, the prognostic value of NHHR in AF recurrence following CA was unclear. In our study, baseline characteristics revealed a negative association between NHHR and age, as well as HDL-C, while a positive correlation was observed between NHHR and the TC, TG, LDL-C, or glucose levels. The results were consistent with those of a previous study [12], which suggested that NHHR might serve as a cardiovascular risk factor. Our study further discussed the association between NHHR and the risk of AF recurrence and revealed the U-shape association between them. In addition, a series of subgroups, including age, sex, AF type, CHA_2_DS_2_-VASc score, LAD, non-PVI lesions, and diabetes mellitus, were utilized as stratification variables, and no significant interaction was observed between NHHR and the stratified variables, suggesting no significant interactions in the association between NHHR and AF recurrence. The results were similar to those of previous studies on the relationship between NHHR and other diseases. Yu et al. demonstrated the presence of a U-shaped association between NHHR and all-cause mortality among participants with diabetes or prediabetes [12, 13]. A U-shaped relationship was observed between NHHR and MACCE incidence in patients with coronary artery disease undergoing percutaneous coronary intervention [13].

The U-shaped relationship between NHHR and AF recurrence risk demonstrated a dual-pathophysiological mechanism, where deviations from the optimal lipid equilibrium (NHHR=2.61) in either direction independently predispose to arrhythmia. Our threshold analysis identified 2.61 as the critical inflection point, with the lowest recurrence risk observed within the range of 2.61–3.26. Below this threshold, each unit increase in the NHHR reduced the AF recurrence risk by 28.5%, whereas above this threshold, every unit increment elevated the risk by 10.2%. The positive NHHR-AF risk association above 2.61 corresponds well with established atherogenic mechanisms. HDL-C possesses many protective effects, such as anti-inflammatory, anti-oxidative stress, and cholesterol efflux, while non-HDL-C has the opposite effects [24–26]. These effects are also the important influencing factors of AF [27]. The inverse relationship observed at the NHHR values below 2.61—where lower NHHR (indicating higher HDL-C dominance) paradoxically increases the AF recurrence risk—mirrors the “cholesterol paradox” phenomenon seen in AF pathogenesis. A nationwide cross-sectional survey suggested that non-HDL-C may also be associated with a lower risk of AF [28]. Patients with AF exhibit 9% higher levels of oxidized non-HDL compared to individuals without AF, independent of traditional risk factors [29]. Oxidative modifications disrupt HDL’s anti-inflammatory properties, converting it into a pro-oxidant mediator that impairs cholesterol efflux capacity, activates inflammasomes, destabilizes cardiomyocyte membrane lipid rafts, thereby increasing arrhythmia risks, such as AF [29, 30].

This study using real-world data demonstrated some noteworthy advantages; RCS was applied to thoroughly assess the potential association and enhance the study’s ability to uncover the true associations between exposure and outcome. Although no significant dose-effective relationship was observed in patients with NHHR above the inflection point, it may be limited by the relatively small sample size. We conducted subgroup analyses and sensitivity analyses to verify the reliability of these results and further indicated the findings were robust. Clinically, our findings extend the “cholesterol paradox” — previously observed in AF pathogenesis—to post-ablation settings. Both extremes of NHHR imbalance independently predispose to arrhythmia recurrence. Notably, the amplified NHHR-AF recurrence association in males and patients aged < 65 years has suggested age- and sex-specific modulation of lipid metabolism pathways [6, 31]. Estrogen’s regulation of lipid synthesis and age-related declines in LDL receptor activity may partially explain these disparities [32]. Despite NHHR’s prognostic value, current lipid-lowering strategies (statins and PCSK9 inhibitors) primarily target CVD prevention rather than AF-specific mechanisms [33]. While statins demonstrate pleiotropic benefits in reducing systemic inflammation and oxidative stress - mechanisms implicated in AF pathogenesis - their direct impact on NHHR dynamics and AF recurrence remains unproven. Our findings aligned with the "cholesterol paradox" phenomenon, where conventional lipid metrics (LDL-C and HDL-C) exhibited bidirectional associations with AF risk [28]. NHHR’s U-shaped relationship with post-ablation AF recurrence outperforms traditional metrics by integrating both atherogenic burden (non-HDL-C) and reverse cholesterol transport efficiency (HDL-C). The absence of NHHR-MACCE association reinforced that AF-specific lipid pathophysiology might operate independent of the atherosclerotic pathways. Thus, NHHR should not override established CVD lipid targets [19].

There are certain limitations in this study. First, this was an observational study; thus, a causal relationship between NHHR and investigated outcomes could not be established. Second, this study adopted the baseline NHHR for analysis and did not further analyze the relationship between the long-term fluctuation of NHHR and AF recurrence, which may limit the ability to assess the therapeutic necessity of NHHR for patients with AF. Third, this was a single-center study with a relatively small sample size, which may limit the generalizability of our findings to other populations. AF recurrence was assessed through scheduled 12-lead ECGs, 24-hour Holter monitoring, and symptom-triggered ECG evaluations. While this surveillance protocol underestimated the recurrence rates by missing asymptomatic or short-lived AF episodes [34], the detection bias was non-differential across the study groups. Future multicenter studies with larger cohorts are warranted to explore the association between NHHR and AF recurrence.

## Conclusion

In summary, this study demonstrated a novel U-shaped association between NHHR and AF recurrence following CA. These findings extend the "cholesterol paradox" to post-ablation settings, where both excessive atherogenic burden (NHHR>2.61) and dysfunctional reverse cholesterol transport (NHHR<2.61) independently predispose to arrhythmia recurrence. Routine monitoring of NHHR may contribute to evaluation of the recurrence risk of AF ablation in patients.

## Data Availability

The data underlying this article will be shared on reasonable request to the corresponding author.

## LIST OF ABBREVIATIONS

AF: atrial fibrillation
CA: catheter ablation
HDL-C: high-density lipoprotein-cholesterol
NHHR: non-HDL-C/HDL-C ratio
MACCE: major adverse cardio- and cerebrovascular events
HR: Hazard ratio
CI: confidence interval
TC: total cholesterol
TG: triglycerides
LDL: low-density lipoprotein
ECG: electrocardiogram
RFA: radiofrequency ablation
LAD: left atrial diameter
PAF: paroxysmal AF
PV: pulmonary vein
PVI: pulmonary vein isolation

## DECLARATIONS

### Ethics approval and consent to participate

Ethical approval was obtained from the Institutional Review Board of the Qilu Honspital (Approval No. [2014]008), adhering to the Declaration of Helsinki principles. Written informed consent was obtained from all participants.

### Consent for publication

Not applicable.

### Availability of data and materials

The data underlying this article will be shared on reasonable request to the corresponding author.

### Authors’ contributions

Mingjie Lin: Conceptualization, Methodology, Formal analysis, Writing – Original Draft, Visualization; Hao Wang: Investigation, Validation, Data curation, Writing – Review & Editing; Nan Yang: Resources, Software, Validation; Tongshuai Chen: Investigation, Data curation, Formal analysis; Wenqiang Han: Methodology, Project administration; Bing Rong: Supervision, Funding acquisition, Writing – Review & Editing; Jingquan Zhong: Conceptualization, Resources, Supervision, Funding acquisition; Kai Zhang: Conceptualization, Supervision, Funding acquisition, Writing – Original Draft, Review & Editing, Final approval.

## Acknowledgements

Not applicable.

